# Effects of Intravenous and Inhaled Anesthetics on the Postoperative Complications for the patients undergoing One Lung Ventilation

**DOI:** 10.1101/2022.04.01.22273288

**Authors:** Jing Yang, Qinghua Huang, Rong Cao, Yu Cui

## Abstract

**Introduction:** With the widely used technique of One Lung Ventilation (OLV) in patients throughout thoracic surgery, it’s unclear whether inhaled or intravenous anesthetics were associated with postoperative complications. The purpose of the current study is to compare the effects of intravenous and inhaled anesthetics on the postoperative complications within the patients suffering OLV.

**Methods:** We searched the related randomized controlled trials in PubMed\EMBASE\Medline and the Cochrane library up to 09\2021.Inclusive criteria were as follows: We included all the randomized controlled trials which compared the effects of intravenous and inhaled anesthetics on the postoperative complications[listed as: (a) major complications; (b)postoperative pulmonary complications (PPCs); (c) postoperative cognitive function (MMSE score); (d) length of hospital stay; (e) 30-days mortality] for the patients undergoing one lung ventilation.

**Results:** Thirteen randomized controlled trials with 2522 patients were included for analysis. Overall, there were no significant differences in the postoperative major complications between inhaled and intravenous anesthetics groups (OR 0.78, 95%CI 0.54 to 1.13, *p*=0.19; I^2^=0%). However, more PPCs were detected in intravenous groups when compared to inhaled groups (OR 0.62, 95%CI 0.44 to 0.87, *p*=0.005; I^2^=37%). Both the postoperative MMSE scores (SMD -1.94, 95%CI -4.87 to 0.99, *p*=0.19; I^2^=100%) and the length of hospital stay (SMD 0.05, 95%CI -0.29 to 0.39, *p*=0.76; I^2^=73%) were comparable between two groups. Besides, the 30-day mortality didn’t differ significantly across groups either (OR 0.79, 95%CI 0.03 to 18, *p*=0.88; I^2^=63%).

**Conclusions:** In patients undergoing OLV, generous anesthesia with inhaled anesthetics could reduce PPCs compared with intravenous anesthetics, but no evident advantages were provided over other major complications, cognitive function, hospital stay or mortality.

## Introduction

According to the literature, approximately 3% surgical patients develop major complications, resulting in 0.4% deaths peri-operatively(1). And lung cancer is the leading reason of cancer-related death in the United States(2). Since one-lung ventilation (OLV) could facilitate the performance of surgery and prevent contamination of the other lung, it has become a necessary technique during thoracic surgery(3). However, OLV could cause ischemia and hypoxia in the nonventilated lung, barotrauma and excessive fluids in the ventilated lung tissue, and alveolar or even systematic inflammatory responses, thus increasing the risk of postoperative complications(4). The incidence of postoperative pulmonary complications (PPCs) is much higher in patients operated with OLV than in those without OLV(5).

Christopher.U et al. found that in cardiac surgery, inhaled anesthesia was associated with major benefits in outcomes and reduced mortality(6). However, Bassi.A(7) and Modolo.NS(3) revealed that few evidence from randomized controlled trials(RCTs) demonstrated significant difference in particular postoperative outcomes between general anesthesia maintained by inhaled and intravenous anesthetics in case of OLV in 2008 and 2013. Later, several RCTs and systematic reviews suggested that inhalation might preserve intraoperative cardiac function, reduce PPCs, attenuate local alveolar inflammatory responses in patients undergoing OLV(8-10).

Since 2013, increasing clinical RCTs have been published to explore the distinct effects of different sedative anesthetics on the major complications in patients with OLV. Therefore, we conducted this meta-analysis to compare the effects of inhaled anesthetics (Sevoflurane or Desflurane) and intravenous anesthetic (Propofol) regarding patients’ postoperative outcomes.

## Methods

We followed the recommendation from the Cochrane Handbook for the Systematic Review(11). And the meta-analysis has been registered on https://www.crd.york.ac.uk/prospero/ with No. CRD420202222856.

### Retrieval strategy

Two authors (JY, QHH) separately searched the Pubmed, Medline, Embase and Cochrane central register for the relevant RCTs from Jan 1st, 2000 to Sep 31st, 2021. We used various combinations of the key words and MeSH terms to perform the search. The search terms were listed as Table 1 and the search was limited to English.

**Table 1.** The specific keywords and MeSH terms during the screening process

|  |  |  |  |
| --- | --- | --- | --- |
| anesthesia, intravenous | anesthetics, inhalation | one lung ventilation | RCT |
| intravenous anesthesia | anesthetic gases | single lung ventilation | Randomized controlled trial |
| intravenous anesthetics | inhalation anesthetics | Ventilation, One-Lung | Controlled clinical trial |
| intravenous anesthetic agent | inhalation anesthesia | Ventilation, Single-Lung | Randomized |
| propofol | inhaled anesthesia | lung separation techniques | Randomly |
| Diprivan | volatile anesthetics | Separation Technique, Lung | Trial |
| Disopropofol | sevoflurane | Technique, Lung Separation |  |
|  | sevorane | Lobectomy |  |
|  | desflurane | thoracic surgery |  |
|  | isoflurane |  |  |
| We used Boolean operator “OR” to search every potentially eligible article that was relevant to Intravenous anesthetics/ Inhaled anesthetics/ One lung ventilation. Then, we used the operator “AND” to combine the above results to accomplish the screening process. |  |  |  |

#### Inclusion criteria

1. Population: Patients (>18 years old) were scheduled for elective thoracic surgery under OLV.
2. Intervention: Patients receiving anesthesia maintained with inhaled anesthetics when OLV.
3. Comparison: Patients receiving intravenous anesthetics like propofol to maintain the anesthesia when OLV.
4. Outcomes:

The primary endpoint was the number of major complications assessed by Clavien-Dindo score (grade III to V) or not (complications needs more intensive treatments including overall cardiac events, myocardial infarction, acute renal failure, hepatic failure, disseminated intravasal coagulation, extrapulmonary infection, gastrointestinal failure, coma).

The secondary endpoints were the number of PPCs (hypoxemia, acute respiratory distress syndrome, pulmonary infiltrates, pneumonia, pleural effusions, atelectasis, pneumothorax, bronchospasm, cardiopulmonary edema, aspiration pneumonitis); scores of Mini-mental State Examination (MMSE), length of hospital stay and 30-day mortality after OLV.

(5) Study: Randomized controlled studies.

Any trials reported the familiar Population, Intervention, Comparison, Studies, and at least one outcome as listed before were included.

#### Exclusion criteria

Duplicated studies, nonhuman or pediatric studies, conference abstracts, studies published before the 2000s, and the studies which data could not be extracted.

### Data extraction

Based on the criteria above, two authors (JY, QHH) sequentially enrolled the trials and extracted the data independently: publication information (first author’s name, publication year), characteristics of participants (sample size, type of surgery, anesthesia induction schemes, duration of OLV and surgery, OLV strategies) and outcomes information. Disagreements over eligibility between the two researchers were resolved by discussion. If necessary, the third researcher (RC) was involved and adjudicated. The data were extracted or calculated from figures using the program Engauge Digitizer 5.1 if necessary (M. Mitchell, Engauge Digitizer, http://digitizer.sourceforge.net). All the extracted data were collected in the standardized Excel file by the two authors, and YC double-checked the accuracy.

### Risk of Bias Assessment and Strength of Evidence

Two reviewers (JY, QHH) independently employed the method recommended by Cochrane Collaboration to assess the methodological quality of included trials. For each trial, the criteria used for quality assessment were random sequence generation, allocation concealment, performance bias, detection bias, attribution bias, reporting bias, and others. Each criterion is classified as “yes”, “no” or “unclear”, and the brief assessment for each trial were classified as three levels (low risk of bias, unclear risk of bias and high risk of bias). The Grading of Recommendations, Assessment, Development and Evaluations approach (GRADEpro; gdt.gradepro.org) was approved to rating the overall quality of evidence for each outcome. In the approach, each outcome began as high-quality evidence, but may be rated down by one or more of five categories of limitations: risk of bias, inconsistency, indirectness, imprecision, and reporting bias. Finally, the approach drew an evident quality of each outcome as low, moderate or high.

### Statistical Analysis

According to DerSimonian and Laird method performed by Review Manager 5.3 (RevMan, The Cochrane Collaboration, Oxford, UK), differences were expressed as risk ratio (RR) with 95% confidence intervals (CI) for dichotomous data, and the differences between continuous data were expressed as mean differences (MDs) or standardized mean differences (SMD) with 95% CI. Due to the limited number and heterogeneity between included studies, the Hartung-Knapp-Sidik-Jonkman (HKSJ) method were further conducted for random effects to the data pooled by less than 5 trials, or the value of heterogeneity was more than 50%. Since Joanna(12) had proved that HKSJ method were outperformed than DerSimonian-Laird method for the meta-analysis among limited number studies or high heterogeneity.

The heterogeneity between pooled studies was represented by I^2^ value, and the criteria to identify whether the combined data being high or low heterogeneity was 50%. On account of the inconsistency of the surgery process, method of anesthesia, time of OLV and the factors which increased the heterogeneity, random effects model was conducted for significant heterogeneity (I^2^>50%, *p* ≤ 0.1). And sensitivity analyses were implied to explore the possible explanation for the high heterogeneity too.

## Results

### Study Identification

The search yielded 1945 publications by the original screening. Based on the inclusion and exclusion criteria, 1319 potentially eligible trials were excluded based on the title or abstract. Within the full text screening we excluded 46 studies (10 articles were not RCT, 19 studies did not match the population criteria, 9 compared intravenous anesthetics to regional anesthesia or other drugs, and 9 studies did not report any outcome as listed before). Finally, we included 13 studies and meta-analyzed the data(13-25). The flow chart was presented in Fig.1.

**Fig 1.**
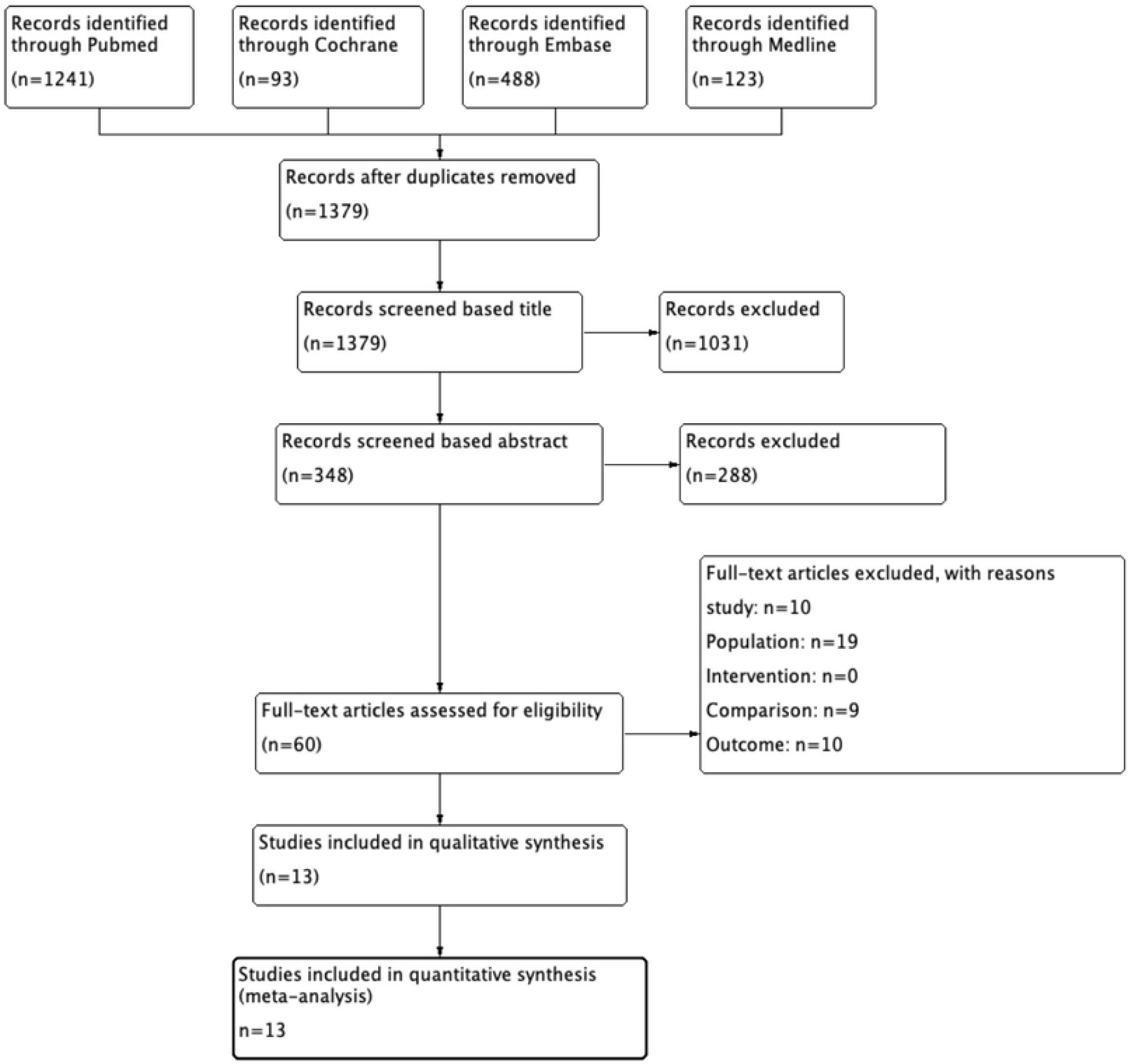
Flow diagram of selecting process

### Study Characteristics and Quality

The main characteristics of the included trials were depicted in Table 2. The 13studies(13-25) included 2522 patients, which published from 2000 to 2021. As showed in Figure.2, 8(13, 15-20) out of 14 studies presented a low risk of random sequence generation and allocation concealment by describing the randomized method in detail. Although 6(14, 17, 21-24) out of 14 failed to report the details of blinding to participants or outcome evaluators, the outcomes were little influenced by the lack of blinding. According to the GRADEpro system, the quality of every outcome were showed in Table 3. The evidence quality of PPCs was high, and the evidence quality of major complications, 30-days mortality and hospital stay were moderate. However, the evidence of MMSE score was low.

**Table 2.** Trial Characteristics

| Trial | Surgery | Intervention(n=1263) |  | Control(n=1259) |  | OLV strategy | Outcome |
| --- | --- | --- | --- | --- | --- | --- | --- |
|  |  | Induction | Maintenance | Induction | Maintenance |  |  |
| Beck-Schimmer<br>2016(13) | Lung surgery | Desflurane(n=230) |  | Propofol(n=230) |  | Vt<sup>c</sup> 4-6 ml/kg<br>FIO<sub>2</sub><sup>d</sup> 0.6-1.0<br>PEEP<sup>e</sup> 5cmH<sub>2</sub>O | Complications<br>(Clavien-Dindo<br>classification),<br>Hospital stay |
|  |  | Etomidate<br>(0.3-0.5mg/kg) | Desflurane<br>(end-tidal<br>concentrations of<br>4.5-7%) | Etomidate<br>(0.3-0.5 mg/kg) | Propofol<br>TCI<sup>b</sup> (2-6ug/ml) |  |  |
| Conno<br>2009(14) | Lung surgery | Sevoflurane (n=27) |  | Propofol(n=27) |  | Vt<sup>c</sup> 6-7ml/kg<br>FiO<sub>2</sub><sup>d</sup> 1.0 | PPCs,<br>Hospital death |
|  |  | Propofol<br>(1.5-2.5mg/kg) | sevoflurane<br>(1 MAC<sup>a</sup>) | Propofol<br>TCI<sup>b</sup>(3-5ug/ml) | Propofol<br>TCI<sup>b</sup> (1 MAC<sup>a</sup>) |  |  |
| Gala<br>2017(15) | Lung<br>resection<br>surgery | Sevoflurane(n=86) |  | Propofol(n=88) |  | Vt<sup>c</sup> 6ml/kg<br>FiO<sub>2</sub><sup>d</sup> 0.6-1.0<br>PEEP<sup>e</sup> 5cmH<sub>2</sub>O | Complications<br>(Clavien-Dindo<br>classification),<br>PPCs,<br>Mortality,<br>Hospital stay |
|  |  | Propofol<br>(2-3mg/kg) | Sevoflurane<br>(BIS<sup>f</sup> 40-60) | Propofol<br>(2-3mg/kg) | Propofol<br>(BIS<sup>f</sup> 40-60) |  |  |
| Egawa<br>2016(16) | Lung surgery | Sevoflurane(n=72) |  | Propofol(n=72) |  | Vt<sup>c</sup> 5-6ml/kg<br>FiO<sub>2</sub><sup>d</sup> 1.0<br>PEEP<sup>e</sup> 4-5cmH<sub>2</sub>O | MMSE score |
|  |  | Propofol<br>(1-2mg/kg) | Sevoflurane<br>(BIS<sup>f</sup> 40-60) | Propofol<br>TCI<sup>b</sup> (3-4ug/ml) | Propofol<br>(BIS<sup>f</sup> 40-60) |  |  |
| Lee<br>2012(17) | Ivor Lewis<br>operation | Sevoflurane(n=24) |  | Propofol(n=24) |  | Vt<sup>c</sup> 6ml/kg<br>FIO<sub>2</sub><sup>d</sup> (achieve<br>oxygen saturation<br>>95%)<br>PEEP<sup>e</sup> 5cmH<sub>2</sub>O | Hospital complications,<br>PPCs,<br>Hospital death,<br>Hospital stay |
|  |  | Thiopental<br>(4-5 mg/kg) | Sevoflurane<br>(end-tidal<br>concentrations of 1-2.5%) | Propofol<br>(BIS<sup>f</sup> 30-50) | Propofol<br>(BIS<sup>f</sup> 30-50) |  |  |
| Li<br>2021(18) | Lung surgery | Sevoflurane(n=169) |  | Propofol(n=167) |  | Vt<sup>c</sup> 6ml/kg<br>FiO<sub>2</sub><sup>d</sup> 0.4-0.5<br>PEEP<sup>e</sup> 5-8cmH<sub>2</sub>O | Complications (Clavien-Dindo classification),<br>PPCs,<br>Death, |
|  |  | Propofol<br>(1.5-2.5mg/kg) | Sevoflurane<br>(BIS<sup>f</sup> 40-60) | Propofol<br>(1.5-2.5mg/kg) | Propofol<br>(BIS<sup>f</sup> 40-60) |  |  |
| Mahmoud | Lung surgery | Isoflurane(n=25) |  | Propofol(n=25) |  | Vt<sup>c</sup> 10ml/kg | PPCs, |
| 2011(19) |  | Propofol<br>(1.5-2 mg/kg) | Isoflurane<br>(1MAC<sup>a</sup>) | Propofol<br>(1.5-2 mg/kg) | Propofol<br>(4-6mg /kg/h) | FiO<sub>2</sub><sup>d</sup> 0.8-1.0<br>PEEP<sup>e</sup> 5cmH<sub>2</sub>O | 30-mortality,<br>Hospital stay |
| Potočnik<br>2014(20) | Thoracic<br>surgery | Sevoflurane(n=17) |  | Propofol(n=19) |  | Vt<sup>c</sup> 4ml/kg<br>FiO<sub>2</sub><sup>d</sup> 0.6-0.7<br>PEEP<sup>e</sup> 3 cmH<sub>2</sub>O | PPCs,<br>Hospital death |
|  |  | Sevoflurane (6<br>%) | Sevoflurane<br>(2-2.5%) | Propofol<br>(1.5-2.0 mg/kg) | Propofol<br>(4-6 mg/kg/h) |  |  |
| Shen<br>2011(21) | Thoracic<br>surgery | Sevoflurane (n=30) |  | Propofol(n=30) |  | Vt<sup>c</sup> 8ml/kg<br>FiO<sub>2</sub><sup>d</sup> 0.6 | MMSE score |
|  |  | Sevoflurane<br>(4-6%) | Sevoflurane<br>(0.8-1.2MAC<sup>a</sup>) | Propofol<br>(1.5-2 mg/kg) | Propofol<br>(6-8mg /kg/h) |  |  |
| Tian<br>2017(22) | Lobectomy | Sevoflurane(n=31) |  | Propofol(n=31) |  | Not reported | Adverse reaction,<br>MMSE score |
|  |  | Sevoflurane<br>(8%) | Sevoflurane<br>(2%) | Propofol<br>(1mg/kg) | Propofol<br>(6mg/kg) |  |  |
| Wang<br>2019(23) | Lung surgery | Sevoflurane(n=32) |  | Propofol(n=26) |  | Vt<sup>c</sup> 8-10ml/kg<br>FiO<sub>2</sub><sup>d</sup> 1.0 | MMSE score |
|  |  | Sevoflurane<br>(6%) | Sevoflurane<br>(1MAC<sup>a</sup>) | Propofol<br>TCI<sup>b</sup> (3ug/kg) | Propofol<br>TCI<sup>b</sup> (4ug/kg) |  |  |
| Xu<br>2014(24) | Open-chest<br>thoracotomy | Sevoflurane(n=20) |  | Propofol(n=20) |  | Vt<sup>c</sup> 8ml/kg<br>FiO<sub>2</sub><sup>d</sup> 1.0 | Complications,<br>PPCs,<br>Hospital death,<br>Hospital stay |
|  |  | Sevoflurane<br>(8%) | Sevoflurane<br>(BIS<sup>f</sup> 40-60) | Propofol<br>TCI<sup>b</sup> (6ug/ml) | Propofol<br>(BIS<sup>f</sup> 40-60) |  |  |
| Yu<br>2017(25) | Thoracic<br>surgery | Sevoflurane(n=500) |  | Propofol(n=500) |  | Vt<sup>c</sup> 8ml/kg | MMSE score |
|  |  | Sevoflurane<br>(2-4%) | Sevoflurane<br>(BIS<sup>f</sup> 45-55) | Propofol<br>(2mg/kg) | Sevoflurane<br>(BIS<sup>f</sup> 45-55) |  |  |

MAC<sup>a</sup>: minimum alveolar concentration; TCI<sup>b</sup>: target controll infusion; Vt<sup>c</sup>: tidal volume; FiO<sub>2</sub><sup>d</sup>: Fraction of inspiration O<sub>2</sub>; PEEP<sup>e</sup>: positive end expiratory pressure; BIS<sup>f</sup>: bispectral index

**Fig 2.**
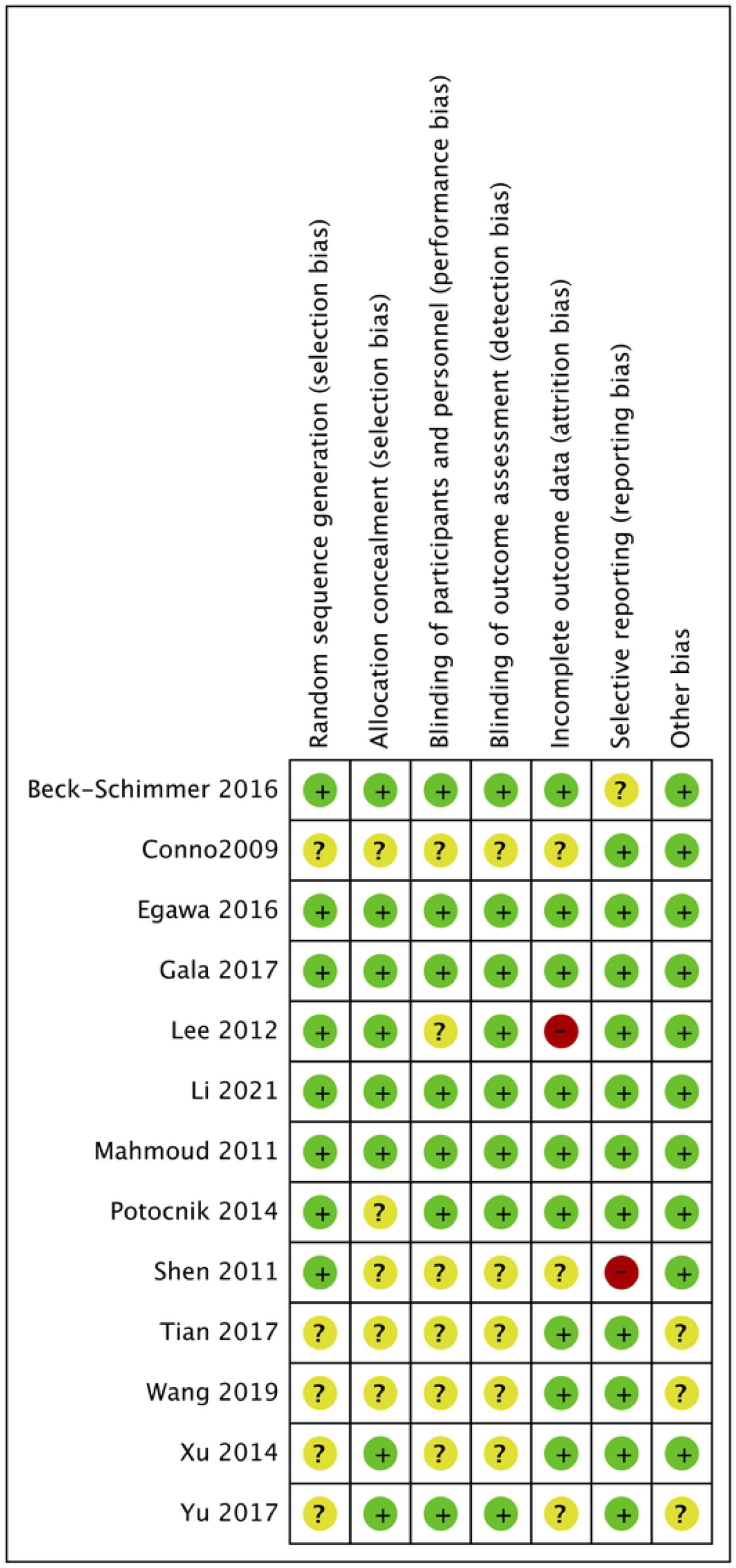
A summary of assessment of risk bias of each RCTs

**Table 3:** The details of GRADE evidence among each outcome

| Certainty assessment |  |  |  |  |  |  | № of patients |  | Effect |  | Certainty | Importance |
| --- | --- | --- | --- | --- | --- | --- | --- | --- | --- | --- | --- | --- |
| № of studies | Study design | Risk of bias | Inconsistency | Indirectness | Imprecision | Other considerations | Inhaled anesthetics | Intravenous anesthetics | Relative (95% CI) | Absolute (95% CI) |  |  |

major complications
| Certainty assessment |  |  |  |  |  |  | № of patients |  | Effect |  | Certainty | Importance |
| --- | --- | --- | --- | --- | --- | --- | --- | --- | --- | --- | --- | --- |
| № of studies | Study design | Risk of bias | Inconsistency | Indirectness | Imprecision | Other considerations | Inhaled anesthetics | Intravenous anesthetics | Relative (95% CI) | Absolute (95% CI) |  |  |
| 5 | randomised trials | not serious | not serious | not serious | serious | none | 60/542 (11.1%) | 74/541 (13.7%) | **OR 0.78**<br>(0.54 to 1.18) | **27 fewer per 1,000**<br>(from 58 fewer to 21 more) | ⊕⊕⊕○<br>Moderate |  |
| **PPCs** |  |  |  |  |  |  |  |  |  |  |  |  |
| 7 | randomised trials | not serious | not serious | not serious | not serious | none | 80/381 (21.0%) | 113/382 (29.6%) | **OR 0.62**<br>(0.44 to 0.87) | **89 fewer per 1,000**<br>(from 140 fewer to 28 fewer) | ⊕⊕⊕⊕<br>High |  |
| **MMSE scores** |  |  |  |  |  |  |  |  |  |  |  |  |
| 5 | randomised trials | serious | serious | not serious | not serious | none | 665 | 659 | - | **SMD 1.94 SD lower**<br>(4.87 lower to 0.99 higher) | ⊕⊕○○<br>Low |  |
| **Mortality (follow-up: 30 days)** |  |  |  |  |  |  |  |  |  |  |  |  |
| 5 | randomised trials | not serious | serious | not serious | not serious | none | 3/335 (0.9%) | 4/335 (1.2%) | **OR 0.79**<br>(0.03 to 18.00) | **2 fewer per 1,000**<br>(from 12 fewer to 167 more) | ⊕⊕⊕○<br>Moderate |  |
| **Length of hospital stay** |  |  |  |  |  |  |  |  |  |  |  |  |
| 5 | randomised trials | not serious | serious | not serious | not serious | none | 385 | 387 | - | **SMD 0.05 higher**<br>(0.29 lower to 0.39 higher) | ⊕⊕⊕○<br>Moderate |  |

#### 1. The primary outcome: major postoperative complications

Five studies(13, 15, 17, 18, 24) assessed the major complications post-surgery including 1083 patients, among which fixed effects model showed that there have no significantly difference between intravenous or inhaled anesthetics groups (OR 0.78, 95%CI 0.54 to 1.13, *p*=0.19; I^2^=0, Fig.3.).

**Fig 3.**
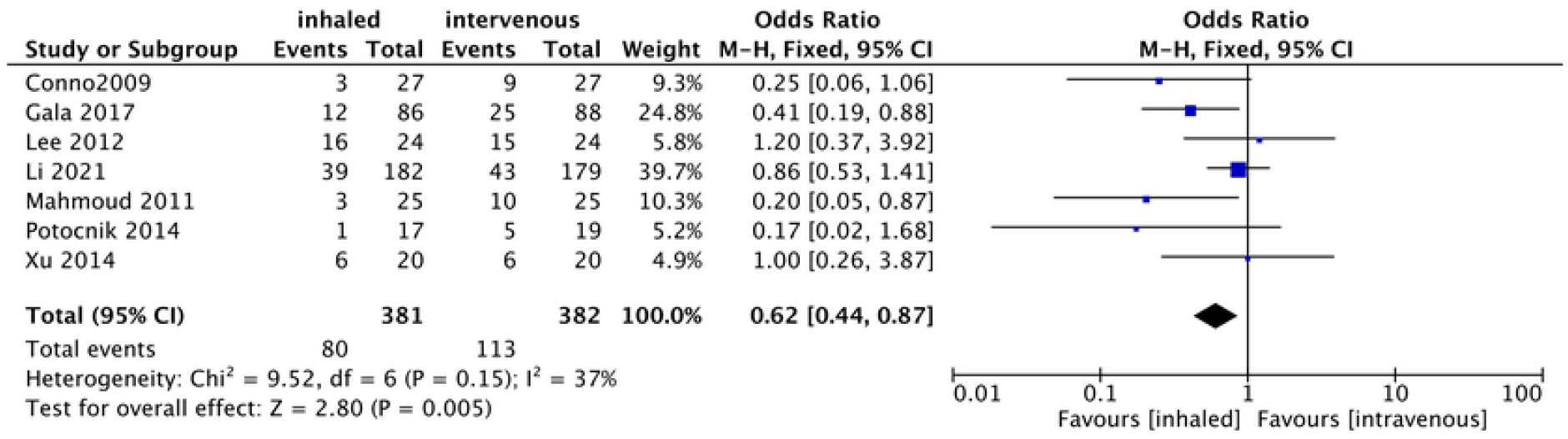
Forest plot for the number of postoperative major complications between inhaled and intravenous groups

#### 2. The second outcomes

##### 2.1 PPCs

Seven RCTs(14, 15, 17-20, 24) compared the effect of intravenous and inhaled anesthetics on PPCs in 763 patients with OLV. The fixed effects model depicted that compared to intravenous anesthetics, inhaled anesthetics significantly reduced the number of patients who had PPCs with low heterogeneity (OR 0.62, 95%CI 0.44 to 0.87, *p*=0.005; I^2^=37%, Fig.4.).

**Fig 4.**
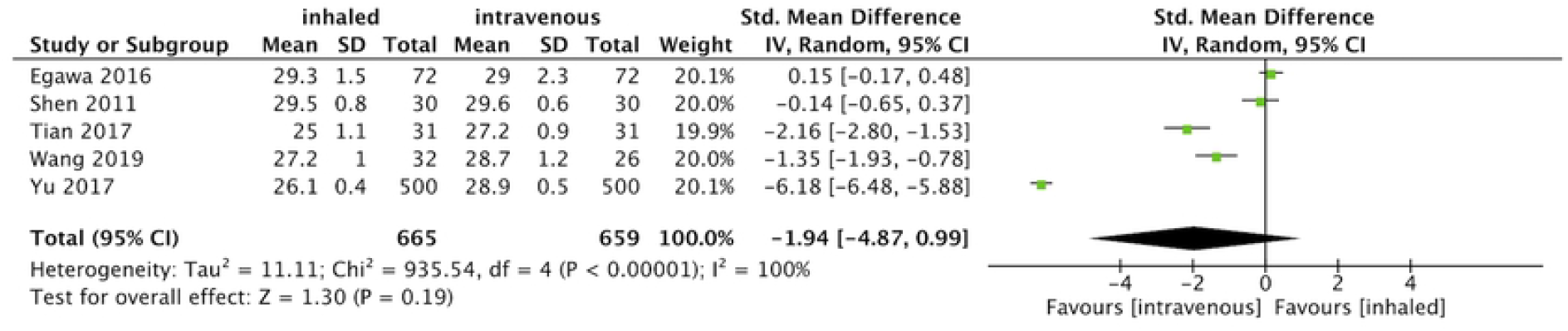
Forest plot for the number of postoperative pulmonary complications between inhaled and intravenous groups

##### 2.2 Postoperative MMSE scores

As Fig.5. showed, five RCTs(16, 21-23, 25) estimated the postoperative cognitive function after OLV through MMSE scores in 1324 patients. The pooled data concluded that anesthetics barely have effect on MMSE scores (SMD -1.94, 95%CI -4.87 to 0.99, *p*=0.19; I^2^=100%). In terms of the extremely high heterogeneity, sensitivity analysis and HKSJ method were further conducted to strength the outcome. Nonetheless, we failed to obtain the original heterogeneity study by removing any individual trials and HKSJ method reached the same conclusion as before (SMD -1.94, 95%CI -5.11 to 1.23, *p*=0.16).

**Fig 5.**
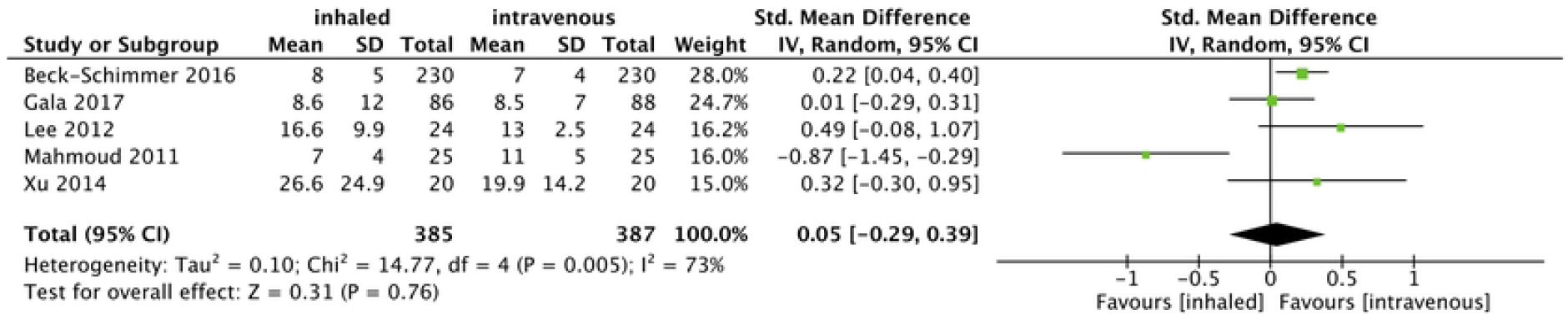
Forest plot for the postoperative MMSE scores between inhaled and intravenous groups

##### 2.3 Length of hospital stay

Data for the length of hospital stay were extracted from five trials(13, 15, 17, 19, 24) with 772 patients. Fig.6. revealed that the types of anesthetics were not associated with significant differences in the length of hospital stay at all (SMD 0.05, 95%CI -0.29 to 0.39, *p*=0.76; I^2^=73%, Fig.6.). Sensitivity analysis detected that Mahmoud 2011(19) contributed to the total heterogeneity. After removing this study, the pooled data outlined that the patients in the intravenous group spent significantly less time in hospital (SMD 0.19; 95 % CI 0.05 to 0.34, *p*= 0.01; I^2^=0) compared with patients in the inhaled group. However, HKSJ method strength the conclusion that anesthetics were not related to the length of hospital stay after excluded the Mahmoud 2011 and depicted the instability of result above (SMD 0.19, 95%CI -0.04 to 0.42, *p*=0.07).

**Fig 6.**
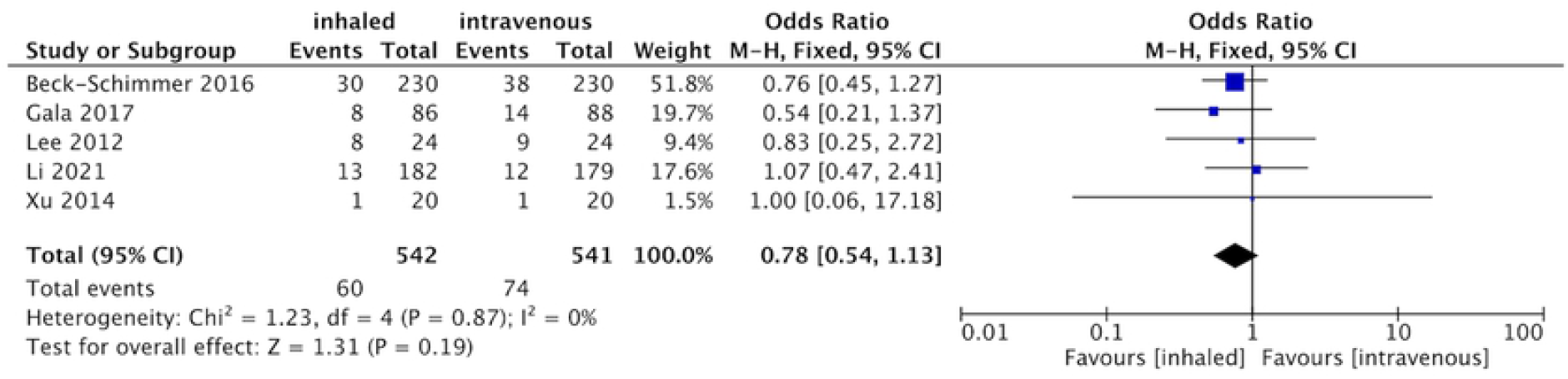
Forest plot for the length of hospital stay between inhaled and intravenous groups

##### 2.4 30-days mortality

Two out of five studies(15, 17-19, 24) which intended to assess the mortality within 30 days after surgery and found that there has no difference in 30-days mortality between two groups (SMD 0.79, 95%CI 0.03 to 18, *p*=0.88; I^2^=63%, Fig.7.).

**Fig 7.**
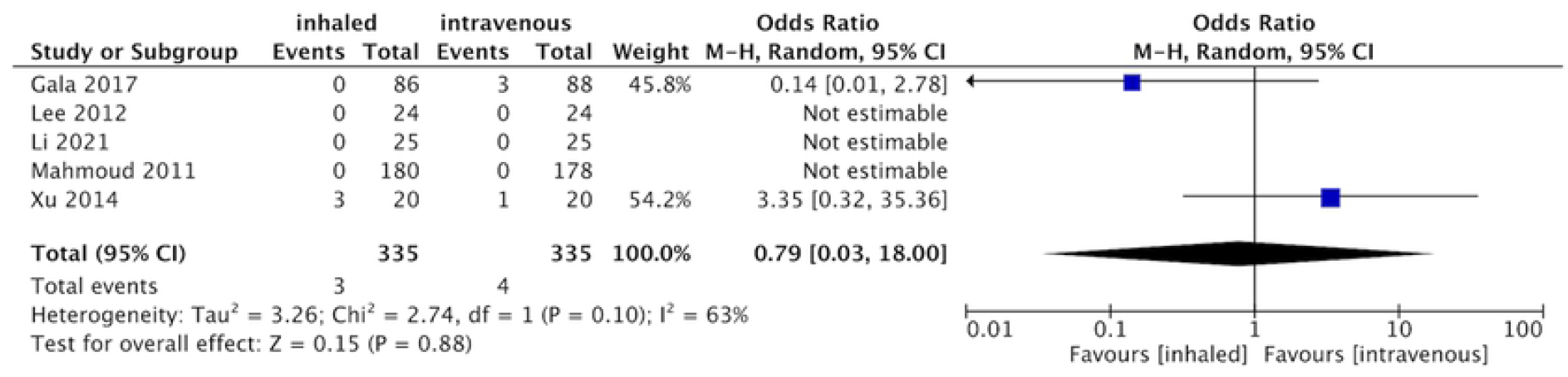
Forest plot for the 30-day mortality between inhaled and intravenous groups

## Discussion

In the present analysis, we included 13 eligible trials(13-25) with 2522 patients who underwent OLV to illustrate that compared to intravenous anesthetics, inhaled anesthetics were associated with fewer risk of PPCs with the high evidence evaluated by GRADEpro system. However, different types of anesthetics had no significantly different effect on the major complications, postoperative MMSE scores, length of hospital stay or 30-days mortality.

According to the meta-analysis accomplished by Uhlig et.al(6), general anesthesia with inhaled anesthetics was associated with the reduction of major complications and mortality in cardiac surgery, which might possibly due to the cardioprotective properties of volatile anesthetics through coronary vasodilation and reduced stress responses(26).Similarly, Uhlig(6) concluded that total inhaled anesthetics seemed to be associated with less benefits in major complications, mortality, length of hospital stay compared to intravenous anesthetics in noncardiac surgery. Even though, studies had demonstrated that the anti-inflammatory effects of volatile anesthetics affected other organs like lungs, brain, kidneys and liver in virto(27-29). In terms of patients receiving noncardiac surgery (including thoracic, vascular and abdominal surgery), major complications, length of hospital stay and mortality might be more relevant to types of surgery, characteristic of patients, different standardized operating procedures rather than types of anesthetics. Thus, the systematic organ protection of inhaled anesthetics was diluted for the patients undergoing OLV that we discussed.

As far as we are concerned, inhaled anesthetics interferes with hypoxic pulmonary vasoconstriction (HPV), and causes hypoxemia when used at a minimum alveolar concentration greater than one when OLV(30). However, Prakash(31) observed that volatile agents had a direct effect on bronchial smooth muscle and contributed to bronchodilation, which acted with lower pressures and greater Cdyn in cases of OLV when compared with propofol. Thus, there are both advantages and disadvantages of inhaled anesthetics on the pulmonary function when OLV.

Regarding to inflammatory responses in vitro(32) or in vivo(33), inhaled anesthetics significantly reduced inflammatory responses to lung injury, contributing to immunomodulatory and organ protective effects. In case of the clinical surgery proceeded with OLV, inhaled anesthetics were found to play anti-inflammatory role by acting on the cytokine responses, ischaemia-reperfusion and oxidative stress(15, 34). Besides, meta-analysis concluded that compared with total intravenous anesthesia, total inhaled anesthesia could reduce the alveolar inflammatory responses but had no significantly effect on the systematic inflammatory responses in the meantime(10), which may contribute to the results that inhaled agents have a benefit effect on the incidence of PPCs rather than the other systematically complications.

Given the report by International Perioperative Neurotoxicity Working Group, few evidence is detected on which particular anesthetic is preferable to postoperative cognitive function in general anesthesia(35). Trials have proved that cereal oxygenation saturation is associated with postoperative cognitive dysfunction(36), and OLV is relevant to a definite reduction in partial pressure of oxygen when compared to baseline as well(37). Moreover, trials revealed that oxygenation index were higher in intravenous anesthetics groups compared to inhaled group within the first 30 minutes after OLV(8). However, in consistent with the Working Group’s Recommendation, we found postoperative cognitive function screening (MMSE scores) were still comparable between two groups after OLV. The reasons may lay in that MMSE screening tool is insufficient to measure cognitive function, and the following time of included trials may be not long enough due to postoperative cognitive dysfunction may last for weeks to months. More importantly, desaturation was rare in all participants, which may offset the different effect of anesthetics on cognitive function.

We came across several limitations through the analysis. First, not all included trials applied Clavien-Dindo score to assess major complications systematically in which complications were analyzed with 0 to 5 different severity grades. Due to limited published articles, to reduce the risk of bias as far as possible, the postoperative events are defined as the events that need more intensive treatments. Second, only two included trials reported the rate of mortality. The mortality is relatively low and affected more by multi-factors than anesthetics alone. Thus, the conclusion may be referenced with consideration. Next, parts of data we acquired was transferred from median/range or graph. Although the method we chose was used commonly, the data was not completely original and may add to the risk of error rate. Finally, we limited the language to English, which may increase the risk of publication bias. Thus, as two Cochrane Meta-analysis recommended(3, 7), if researchers doubted the prognosis of different anesthetics when OLV, they should design and carry out more high-quality and large-scaled trials to assess the standardized outcomes in the future.

## Conclusion

For the patients suffering OLV, compared to intravenous anesthetics, inhaled anesthetics showed significantly protective effects on the PPCs, but not on the postoperative major complications, cognitive function, hospital stay or 30-days mortality either. Further studies are required to test and verify the conclusion.

## Key messages

1. Comparing to propofol, inhaled anesthetics provided protective effects over postoperative pulmonary complications for the patients undergoing OLV.
2. No significantly advantages were observed on the other major complications, cognitive function, hospital stay or 30-days mortality between different anesthetics when OLV.

## Data Availability

All relevant data are within the manuscript and its Supporting Information files.

## Authors’ contributions

Conceptualization: Jing Yang, Qing-Hua Huang, Yu Cui. Data retrieval: Jing Yang, Qing-Hua Huang.

Project administration: Yu Cui. Supervision: Rong Cao.

Validation: Rong Cao.

Writing – original draft: Jing Yang, Qing-Hua Huang. Writing – review and editing: Rong Cao, Yu Cui.

All authors read and approved the final version of the manuscript.

### Abbreviations

OLV: one lung ventilation
PPCs: postoperative pulmonary complications
MMSE: Mini-mental State Examination.

## REFERENCEUncategorized References

1. Weiser TG, Regenbogen SE, Thompson KD, Haynes AB, Lipsitz SR, Berry WR, et al. An estimation of the global volume of surgery: a modelling strategy based on available data. Lancet. 2008;372(9633):139–44.

2. Jemal A, Center MM, DeSantis C, Ward EM. Global patterns of cancer incidence and mortality rates and trends. Cancer Epidemiol Biomarkers Prev. 2010;19(8):1893–907.

3. Módolo NS, Módolo MP, Marton MA, Volpato E, Monteiro Arantes V, do Nascimento Junior P, et al. Intravenous versus inhalation anaesthesia for one-lung ventilation. Cochrane Database Syst Rev. 2013;2013(7):Cd006313.

4. Bernasconi F, Piccioni F. One-lung ventilation for thoracic surgery: current perspectives. Tumori. 2017;103(6):495–503.

5. Canet J, Gallart L, Gomar C, Paluzie G, Valles J, Castillo J, et al. Prediction of postoperative pulmonary complications in a population-based surgical cohort. Anesthesiology. 2010;113(6):1338–50.

6. Uhlig C, Bluth T, Schwarz K, Deckert S, Heinrich L, De Hert S, et al. Effects of Volatile Anesthetics on Mortality and Postoperative Pulmonary and Other Complications in Patients Undergoing Surgery: A Systematic Review and Meta-analysis. Anesthesiology. 2016;124(6):1230–45.

7. Bassi A, Milani WR, El Dib R, Matos D. Intravenous versus inhalation anaesthesia for one-lung ventilation. Cochrane Database Syst Rev. 2008(2):Cd006313.

8. Pang QY, An R, Liu HL. Effects of inhalation and intravenous anesthesia on intraoperative cardiopulmonary function and postoperative complications in patients undergoing thoracic surgery. Minerva Anestesiol. 2018;84(11):1287–97.

9. Sun B, Wang J, Bo L, Zang Y, Gu H, Li J, et al. Effects of volatile vs. propofol-based intravenous anesthetics on the alveolar inflammatory responses to one-lung ventilation: a meta-analysis of randomized controlled trials. J Anesth. 2015;29(4):570–9.

10. Yuan JL, Kang K, Li B, Lu J, Miao MR, Kang X, et al. The Effects of Sevoflurane vs. Propofol for Inflammatory Responses in Patients Undergoing Lung Resection: A Meta-Analysis of Randomized Controlled Trials. Front Surg. 2021;8:692734.

11. Moher D, Liberati A, Tetzlaff J, Altman DG, Group P. Preferred reporting items for systematic reviews and meta-analyses: the PRISMA statement. BMJ. 2009;339:b2535.

12. IntHout J, Ioannidis JP, Borm GF. The Hartung-Knapp-Sidik-Jonkman method for random effects meta-analysis is straightforward and considerably outperforms the standard DerSimonian-Laird method. BMC Med Res Methodol. 2014;14:25.

13. Beck-Schimmer B, Bonvini JM, Braun J, Seeberger M, Neff TA, Risch TJ, et al. Which Anesthesia Regimen Is Best to Reduce Morbidity and Mortality in Lung Surgery?: A Multicenter Randomized Controlled Trial. Anesthesiology. 2016;125(2):313–21.

14. De Conno E, Steurer MP, Wittlinger M, Zalunardo MP, Weder W, Schneiter D, et al. Anesthetic-induced improvement of the inflammatory response to one-lung ventilation. Anesthesiology. 2009;110(6):1316–26.

15. de la Gala F, Piñeiro P, Reyes A, Vara E, Olmedilla L, Cruz P, et al. Postoperative pulmonary complications, pulmonary and systemic inflammatory responses after lung resection surgery with prolonged one-lung ventilation. Randomized controlled trial comparing intravenous and inhalational anaesthesia. Br J Anaesth. 2017;119(4):655–63.

16. Egawa J, Inoue S, Nishiwada T, Tojo T, Kimura M, Kawaguchi T, et al. Effects of anesthetics on early postoperative cognitive outcome and intraoperative cerebral oxygen balance in patients undergoing lung surgery: a randomized clinical trial. Can J Anaesth. 2016;63(10):1161–9.

17. Lee JJ, Kim GH, Kim JA, Yang M, Ahn HJ, Sim WS, et al. Comparison of pulmonary morbidity using sevoflurane or propofol-remifentanil anesthesia in an Ivor Lewis operation. J Cardiothorac Vasc Anesth. 2012;26(5):857–62.

18. Li XF, Hu JR, Wu Y, Chen Y, Zhang MQ, Yu H. Comparative Effect of Propofol and Volatile Anesthetics on Postoperative Pulmonary Complications After Lung Resection Surgery: a Randomized Clinical Trial. Anesthesia and analgesia. 2021.

19. Mahmoud K, Ammar A. Immunomodulatory Effects of Anesthetics during Thoracic Surgery. Anesthesiol Res Pract. 2011;2011:317410.

20. Potočnik I, Novak Janković V, Šostarič M, Jerin A, Štupnik T, Skitek M, et al. Antiinflammatory effect of sevoflurane in open lung surgery with one-lung ventilation. Croat Med J. 2014;55(6):628–37.

21. Shen YF, Wu JX, Xu MY. Effects of anesthesia with propofol and sevoflurane on postoperative cognitive function of elderly patients undergoing thoracic surgery. Journal of shanghai jiaotong university (medical science). 2011;31(3):322–5.

22. Tian HT, Duan XH, Yang YF, Wang Y, Bai QL, Zhang X. Effects of propofol or sevoflurane anesthesia on the perioperative inflammatory response, pulmonary function and cognitive function in patients receiving lung cancer resection. Eur Rev Med Pharmacol Sci. 2017;21(23):5515–22.

23. Wang G, Liu J, Gao J, Zheng X. Comparison of the effects of sevoflurane and propofol anesthesia on pulmonary function, MMP-9 and postoperative cognition in patients receiving lung cancer resection. Oncol Lett. 2019;17(3):3399–405.

24. Xu WY, Wang N, Xu HT, Yuan HB, Sun HJ, Dun CL, et al. Effects of sevoflurane and propofol on right ventricular function and pulmonary circulation in patients undergone esophagectomy. International journal of clinical and experimental pathology. 2014;7(1):272–9.

25. Yu W. Anesthesia with propofol and sevoflurane on postoperative cognitive function of elderly patients undergoing general thoracic surgery. Pak J Pharm Sci. 2017;30(3(Special)):1107–10.

26. Liu KX, Xia Z. Potential synergy of antioxidant N-acetylcysteine and insulin in restoring sevoflurane postconditioning cardioprotection in diabetes. Anesthesiology. 2012;116(2):488–9; author reply 9-90.

27. Yang Q, Dong H, Deng J, Wang Q, Ye R, Li X, et al. Sevoflurane preconditioning induces neuroprotection through reactive oxygen species-mediated up-regulation of antioxidant enzymes in rats. Anesth Analg. 2011;112(4):931–7.

28. Kim M, Park SW, Kim M, D’Agati VD, Lee HT. Isoflurane activates intestinal sphingosine kinase to protect against bilateral nephrectomy-induced liver and intestine dysfunction. Am J Physiol Renal Physiol. 2011;300(1):F167–76.

29. Kim M, Ham A, Kim JY, Brown KM, D’Agati VD, Lee HT. The volatile anesthetic isoflurane induces ecto-5’-nucleotidase (CD73) to protect against renal ischemia and reperfusion injury. Kidney Int. 2013;84(1):90–103.

30. Lederman D, Easwar J, Feldman J, Shapiro V. Anesthetic considerations for lung resection: preoperative assessment, intraoperative challenges and postoperative analgesia. Ann Transl Med. 2019;7(15):356.

31. Prakash YS, Iyanoye A, Ay B, Sieck GC, Pabelick CM. Store-operated Ca2+ influx in airway smooth muscle: Interactions between volatile anesthetic and cyclic nucleotide effects. Anesthesiology. 2006;105(5):976–83.

32. Yue T, Roth Z’graggen B, Blumenthal S, Neff SB, Reyes L, Booy C, et al. Postconditioning with a volatile anaesthetic in alveolar epithelial cells in vitro. Eur Respir J. 2008;31(1):118–25.

33. Reutershan J, Chang D, Hayes JK, Ley K. Protective effects of isoflurane pretreatment in endotoxin-induced lung injury. Anesthesiology. 2006;104(3):511–7.

34. Schilling T, Kozian A, Senturk M, Huth C, Reinhold A, Hedenstierna G, et al. Effects of volatile and intravenous anesthesia on the alveolar and systemic inflammatory response in thoracic surgical patients. Anesthesiology. 2011;115(1):65–74.

35. Miles Berger KJS, Charles H. Brown IV, Stacie G. Deiner, Robert A. Whittington, Roderic G. Eckenhoff, and for the Perioperative Neurotoxicity Working Group. Best Practices for Postoperative Brain Health: Recommendations From the Fifth International Perioperative Neurotoxicity Working Group. Anesth Analg. 2018. 1406–13 p.

36. Kim J, Shim JK, Song JW, Kim EK, Kwak YL. Postoperative Cognitive Dysfunction and the Change of Regional Cerebral Oxygen Saturation in Elderly Patients Undergoing Spinal Surgery. Anesth Analg. 2016;123(2):436–44.

37. Tanaka N, Katoh RI, Yamamoto M, Hoshino K, Morimoto Y, Ito YM, et al. Changes in cerebral oxygen saturation during one-lung ventilation determined using spatially resolved spectroscopy and contributing factors. J Clin Anesth. 2020;59:99–100.

